# The utility of wearable headband electroencephalography and pulse photoplethysmography to assess cortical hyperarousal in individuals with stress-related mental disorders

**DOI:** 10.1101/2023.06.12.23291277

**Authors:** Borbala Blaskovich, Esteban Bullón-Tarrasó, Dorothee Pöhlchen, Alexandros Manafis, Hannah Neumayer, Luciana Besedovsky, Tanja Brückl, BeCOME Working Group, Peter Simor, Florian P. Binder, Victor I. Spoormaker

## Abstract

**Study Objectives:** Several stress-related mental disorders are characterized by disturbed sleep, but objective sleep biomarkers are not routinely examined in psychiatric patients. We examined the use of wearable-based sleep biomarkers in a psychiatric sample with headband electroencephalography (EEG) including pulse photoplethysmography (PPG), with an additional focus on microstructural elements as especially the shift from low to high frequencies appears relevant for several stress-related mental disorders (cortical hyperarousal).

**Methods:** We acquired 483 nights and could analyze 372 nights of sufficient quality from 83 healthy participants and those with a confirmed stress-related mental disorder (anxiety-affective spectrum). We analyzed the data with respect to macrostructural and microstructural characteristics according to the newly described spectral slope fitting over the whole frequency spectrum.

**Results:** The headbands were accepted well by patients and the data quality was sufficient for most nights. The macrostructural analyses revealed trends for significance regarding sleep continuity but not sleep depth variables. The spectral analyses yielded no between-group differences except for a group × age interaction, with the normal age-related decline in the low versus high frequency power ratio flattening in the patient group. PPG analyses showed that the mean heart rate was higher in the patient group in pre-sleep epochs, a difference that reduced during sleep and dissipated at wakefulness.

**Conclusions:** Wearable devices that record EEG and/or PPG could be used over multiple nights to assess relevant markers such as sleep fragmentation, cortical hyperarousal, and sympathetic drive throughout the sleep–wake cycle in patients with stress-related mental disorders.

## Introduction

Several stress-related mental disorders are characterized by disturbed sleep. Major depressive disorder has insomnia as one of the core symptoms in its diagnosis [1], with polysomnographic studies revealing reduced sleep continuity, sleep depth, and alterations in rapid eye movement (REM) sleep characteristics such as duration, latency, and the density of eye movements [2]. Moreover, several studies have noted subjective and objective sleep difficulties in anxiety disorders [2,3], whereas post-traumatic stress disorder (PTSD) has subjectively reported insomnia and nightmares in its diagnosis [1], with similar objective alterations in sleep continuity and depth and REM sleep characteristics dependent on age groups [4] and other confounds [5]. Additionally, at the microstructural level, the reduction of power in the lower (delta) frequencies[6,7] and, potentially, a shift from low to high spectral frequencies (also referred to as cortical hyperarousal) [8] appear relevant for several stress-related mental disorders.

Disrupted sleep therefore appears to express partial dysfunction in deep-brain circuitry associated with both sleep and emotion regulation. This includes but is not limited to limbic and paralimbic circuitry, the hypothalamus, and multiple arousal-related brainstem regions [9,10]. Moreover, several sleep disorders such as sleep disordered breathing and restless legs / periodic limb movement disorder are more prevalent in psychiatric samples than in the general population, indicative of these sleep disorders being a risk factor to develop mental disorders (or increased symptomatology) after exposure to chronic stressors or traumatic events [11].

This is of importance for biomarker search in psychiatry because no biomarkers with clinical utility have yet been identified for stress-related mental disorders, whereas sleep can provide a multitude of potentially relevant macro- and microstructural biomarkers of interest, including cortical hyperarousal. However, due to limited availability of sleep laboratories in psychiatric clinics (or practices), sleep is not routinely assessed in psychiatric patients, except for in a subjective manner by questionnaires or clinical interviews. Instead of objective sleep quality, several sleep disorders are diagnosed exclusively based on subjective sleep quality, such as insomnia and nightmares [1].

Furthermore, these objective measurements in laboratory/hospital settings are scarce and usually limited to one or two assessment nights. The ecological validity of such laboratory settings is questionable and may not veridically capture the sleep complaints that participants experience in their home environment. In fact, sleep complaints such as nightmares in people with PTSD are rarely observed in the laboratory [12] or even when measured with classic polysomnography in their clinic beds [13], which is in sharp contrast with self-reports. Studies employing multiple nights in the home environment have been more successful in, for example, capturing nightmares [12,14], indicating that more recording nights are needed to adapt to measurement bias and to capture natural sleep profiles. This may be why macrostructural elements generally do not show such impressive effect sizes in between-group comparisons of psychiatric patients with healthy participants.

The availability of wrist wearables that incorporate heart rate and headband electroencephalography (EEG) in the home environment [15] now opens up the possibility to measure such sleep biomarkers in clinical samples [16,17], with wristwatch wearables showing decent accuracy in sleep versus wake classifications in clinical samples [18], without the same problems as actigraphy alone (low specificity for sleep [19]) and promising sleep staging in healthy participants [20]. Headband EEG provides hypnogram data with considerable overlap with standard polysomnography, except for some over- or underestimations of wakefulness dependent on the device [21].

We examined the use of wearable sleep biomarkers in a psychiatric sample with headband EEG including PPG, with an additional focus on microstructural elements such as cortical hyperarousal. Our hypotheses were that, in line with previous research, we would observe some macrostructural differences between healthy controls and psychiatric patients of moderate size (reduced sleep continuity and depth). With more temporally fine-grained analyses relating to spectral power and heart rate (variability), we anticipated further unhealthy sleep characteristics to manifest in a reduced proportion of low versus high frequency power in the EEG in non-rapid eye movement (NREM) sleep, as well as increased heart rate and reduced heart rate variability throughout sleep in individuals with a confirmed stress-related mental disorder.

## Methods and Materials

### Participants

Participants were part of a larger transdiagnostic study currently running at the Max Planck Institute of Psychiatry called the Biological Classification of Mental Disorders (BeCOME) study (registered on ClinicalTrials.gov: <u>NCT03984084</u>). Unmedicated outpatients with stress-related mental disorders (primarily affective and anxiety disorders) and healthy controls between the ages of 18 and 65 years are recruited at the intake procedure at the clinic. Clinical diagnoses of such stress-related mental disorders are verified with the Munich-Composite International Diagnostic Interview (DIAX / M-CIDI), and together with in-depth physiological, imaging, and omics assessment biologically serve the overarching goal of identifying biologically informed subtypes of patients in a transdiagnostic manner. As part of this study protocol, participants are asked to take an EEG headband home at the first appointment and wear it for 1–2 weeks before returning for two days of extensive testing.

To date, 483 nights have been collected, with the number of nights per participant varying between 1 and 17 with a mean of 4.5 nights (SD = 2.7). Around 85% of participants slept 2–9 consecutive nights with the headband. In all of the following analyses, only nights with a minimum record quality of 70% (percentage of the night recording that can be scored according to the automated Dreem analysis software) were included, which resulted in 372 nights (Figure 1).

**Figure 1.**
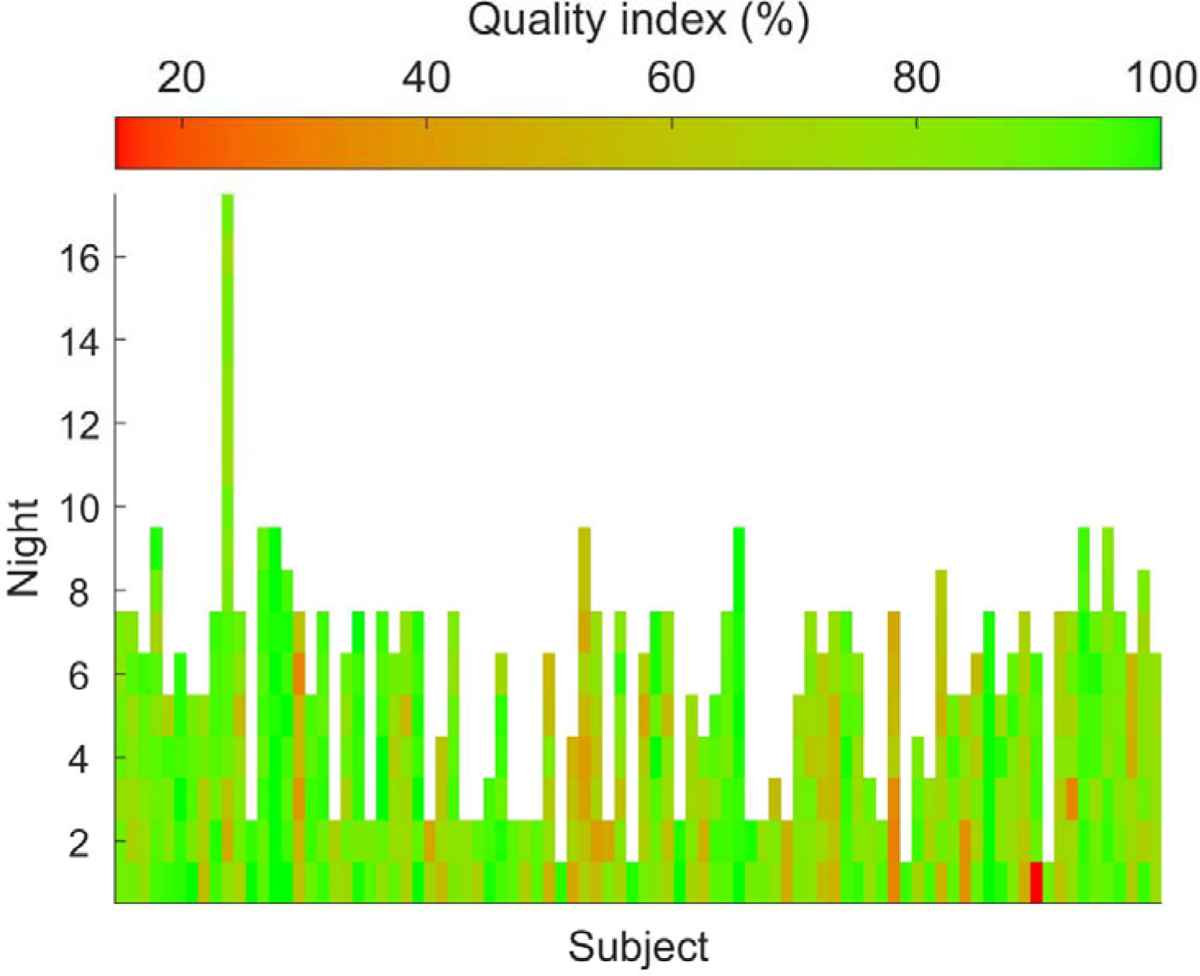
Dreem record quality of all recorded nights (in a consecutive manner) for each participant. Record quality index shows what percentage of the night is still scorable on at least one channel during one recording. On this heatmap, green represents a high record quality and red a low record quality. The first quality assurance step was based on these record quality measurements.

One participant had to be excluded because of missing data (no age information). Finally, 371 nights from 83 participants were included in our analyses (see Table 1 for distributions) with an average age of 33.5 years (*M*_patient_ = 33.9, SD_patient_ = 14.7, *M*_control_ = 33.1, SD_control_ = 13.7).

**Table 1.** Distribution of the included number of participants and nights after exclusion and quality check.

|  | Participant (#) |  |  | Night (#) |  |  |
| --- | --- | --- | --- | --- | --- | --- |
|  | Female | Male | Sum | Female | Male | Sum |
| **Patient** | 33 | 14 | 47 | 134 | 77 | 211 |
| **Control** | 26 | 10 | 36 | 107 | 53 | 160 |
| **Sum** | 59 | 24 | **83** | 241 | 130 | **371** |

All participants provided a written informed consent and had been reimbursed for their participation. The study protocol was in line with the Declaration of Helsinki [22] and was approved by the local ethics committee (reference number: 350 – 14).

### Data acquisition

Sleep data was collected with the Dreem 2 headband (Dreem, Paris, France), a wireless EEG headband with six conductive high-consistency silicone rubber (dry) EEG sensors at locations Fp1, Fp2(ground), F7, F8, O1, O2 with sampling frequency of 250 Hz. These sensors are referenced to each other resulting in the following seven channels: F7-O1, F8-O2, Fp1-F8, F8-F7, Fp1-O1, Fp1-O2, Fp1-F7. Each headband has a three-dimensional axis linear accelerometer measuring movement, position, and breathing frequency and a pulse sensor in LED reflective solution measuring heart rate (both with a sampling rate of 50 Hz). The headbands were handed out on the first day of the study. After a short introduction to the wearable device, nightly recordings were started and finished by the participants (by pushing the start/stop button on the headband). Around 1–2 weeks later, they returned the headbands to the laboratory on the second day of the two-day testing period, and data was uploaded to the server via Wi-Fi.

### Sleep macrostructure and spectral analysis

Sleep stages and conventional macrostructural parameters were scored and calculated by Dreem’s own automatic algorithm [23]. To verify the validity of this algorithm, half of our recordings were manually examined and, where necessary, rescored by two trained scorers. Sensitivity, specificity, and accuracy were calculated for each traditional stage separately (see Supplementary Table S1), resulting in an overall accuracy of 92%. For us, this sufficed to use the traditional macrostructure, sleep-related variables per night, which were summarized for each participant by taking the median value over all recorded nights. NREM stage 1 was left out from the calculation because only 0.6% of the scored epochs belonged to this category based on Dreem’s automatic scoring.

For spectral analyses, raw data was first bandpass filtered between 1 Hz and 40 Hz with a fourth order for the high-pass and a tenth order for the low-pass filter. The data was additionally filtered with a notch filter (tenth order with 48–52 Hz cut-off, to suppress line noise, which was so strong in a few nights that even bandpass filtering could not completely remove it). After filtering, artifact correction was performed by in-house MATLAB (version R2021b, MathWorks Inc., Natick, MA) scripts. This comprised an automated algorithm to identify two of the most common artifact categories characteristic of Dreem 2 recordings (for details, see Supplementary Figure S1). The first category can be described as large amplitude artifacts, most likely the result of movement, electrode displacement, reduction in impedance, or device disconnection. To identify these artifacts, a sleep phase specific, flexible threshold was computed for each recording as a certain quantile of the distribution of all amplitudes in the given sleep phase. The quantile was chosen according to the characteristics of each phase. For NREM stage 2, the 99.5th quantile was chosen, the highest among all sleep phases, to avoid the detection of sleep spindles. Conversely, the quantile was lowered to the 95th for the wake phase, as the proportion of artifactual signal was believed to be much higher in this phase. Lastly, for slow-wave sleep (SWS) and REM, the 99th quantile was sufficient. Note that we did not analyze REM sleep as eye movement detection algorithms would first have to be developed. The filtered signal was divided into 4 s epochs with a 50% overlap, and every epoch that surpassed the given threshold at any of its time points was marked as an artifact. The second category comprised cardiac artifacts. Due to the position and nature of the dry sensors on the headband, the electrodes on the frontal band often shifted onto one of the vessels on the forehead and the signal became tainted with periodically appearing artifactual waves. Since pulse artifacts are time-locked to the heartbeat of the participant, information obtained from the PPG sensor was used to better identify these artifacts. First, PPG peaks were detected and mean RR intervals were calculated for each 12 s 50% overlapping window of the PPG signal. Second, cross correlations (i.e., the correlation of the signal with itself) were calculated in the same epoch basis on the EEG channels for different temporal phase delays. Even though cross correlations always show the maximum peak for lag time 0, in the case of cardiac artifacts, further periodical peaks from the alignment of the pulses in the EEG may appear. An epoch was marked as an artifact when the second largest peak in the cross correlation was located within a lag of +/−100 ms around the pulse detected on the PPG and the amplitude of this peak was higher than 0.4 (maximum peak = 1). Artifacts were marked on each channel separately, and 4 s epochs containing an artifact on any of the channels were excluded from all spectral analyses. This conservative combined method resulted in an average of 59% of clean data in the control group and 53% of clean data in the patient group to be used for further spectral analyses (see performance of individual and combined filters in Supplementary Figure S2).

Artifact-free, 50% overlapping, 4 s epochs were Hanning-tapered and short-time fast Fourier transformed to extract spectral features. To avoid redundancy and increase statistical power by reducing the number of variables, specific spectral features were calculated based on the recommendations of Bódizs and colleagues [24]. For each night, spectral power was calculated between 1 Hz and 30 Hz (sampling frequency of the device was 250 Hz). NREM spectral power was averaged throughout the night and log-log transformed to allow the fitting of a linear regression. Low-frequency spectral power values were interpolated to ensure equidistant frequency steps, and a linear fit was performed on each night in the 2–30 Hz range. However, during NREM sleep, alpha and sigma (sleep spindle) frequencies could entail large peaks, which may confound the fit of the linear regression. Therefore, these frequency bands (6–18 Hz) were excluded from the fitting, as recommended by [24]. Furthermore, from the frequency range of 9–18 Hz, maximum spectral peak amplitude and frequency was extracted to characterize spindle activity during the night. To verify whether our data acquired with the EEG headband showed similar quantitative spectral power profiles to those in the literature, we attempted to replicate the results published by Bódizs and colleagues [24]. Similar to Bódizs et al., our sample showed an age-related significant decline in spectral peak amplitude on most of the channels (−.216 < *r* < −.202, .04 < *p* < .06) and no age-related difference in spindle spectral peak frequency (−.096 < *r* < −.075, .388 < *p* < .503). However, in contrast to the previous study, in our sample there was no significant flattening of NREM spectral slope with age on any of the channels (.022 < *r* < .097, .381 < *p* < .845). Correlations between these three variables and age on the two long-range channels (F7-O1, F8-O2) are presented in Supplementary Figure S3.

Finally, the overnight ascending gradient of NREM slopes was calculated per night. First, the above described linear fitting process was run on every individual 4 s epoch, resulting in as many slope and intercept values as the (artifact-corrected) night had epochs. Thereafter, a linear regression was fitted to the slopes and intercepts of all NREM epochs (see Figure 2) to acquire information about the gradient change and dynamicity of NREM spectral components throughout the whole night. The overnight ascending gradient was computed for each night and summarized per participant with the median value over nights. Outliers +/−3 times the median absolute deviation were excluded from the analysis for each channel separately.

**Figure 2.**
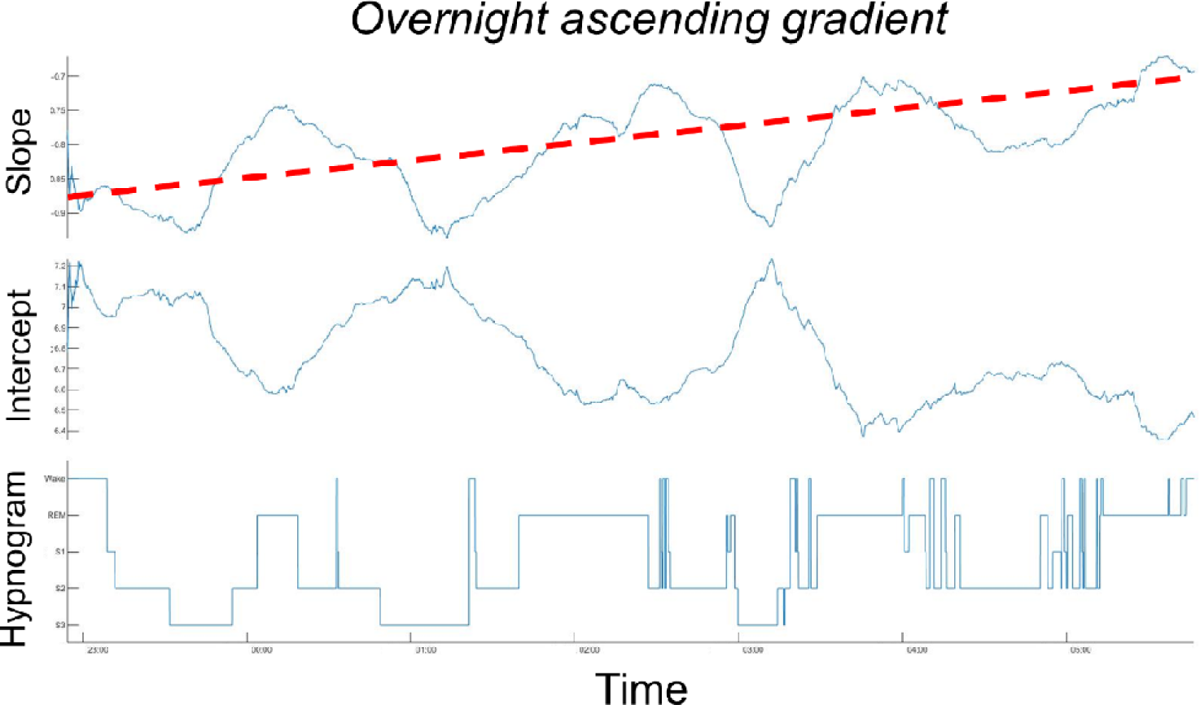
Example of the overnight ascending gradient over one night of a participant. The slope and intercept values of each 4 s epoch are presented on the first two subplots for comparison with the hypnogram of the same night represented on the third subplot. The overnight dynamicity of slopes and intercepts are further smoothed by a 2000 s moving mean for better visualization. The red dashed line represents the linear regression of the NREM slopes (i.e., the overnight ascending gradient). Slopes and intercepts were highly anti-correlated throughout all nights.

### Photoplethysmography analysis

PPG data was high-pass filtered at 0.2 Hz (eighth-order filter) smoothed with a moving mean window of 0.2 s and z-transformed to standardize the variance.

To find the peaks in the PPG data, we used the findpeaks function in MATLAB with a peak width between 0.1 s and 3 s, and optimized the peak location by finding maxima within a window of 0.2 s around the original peaks. If no peak was detected during 2 s with this method, we assumed there was a missing peak. Then, the findpeaks function was re-run in that window on the second derivative of the signal, after filtering the first derivative with a low-pass filter at 1 Hz, to better detect change points at a temporal resolution that was not too high. After this, the difference between consecutive peaks is the RR interval, and the root mean square of these successive differences (RMSSD) was computed for 20 s epochs.

### Statistical analyses

Statistical analyses were performed with MATLAB (version 9.11.0.1769968 (R2021b)) and JASP [25]. Normality of the variables was assessed by a Shapiro–Wilk test, as well as skewness and kurtosis of data distribution. Macrostructural differences between patients and controls were calculated by independent samples *t*-tests or Mann–Whitney *U* tests (if the assumption of normality was violated). To control for multiple comparisons, the Benjamini–Hochberg procedure was used to estimate false discovery rate (FDR). Group dependent spectral slope differences were examined by an analysis of covariance (ANCOVA) with age as a covariate. Differences related to mean heart rate and heart rate variability (RMSSD) across wakefulness and sleep stages were examined by repeated measures analysis of variance (rmANOVA) models. Original (uncorrected) degrees of freedom and corrected *p* values (if applicable) are reported.

## Results

### Sleep architecture

Table 2 summarizes all sleep fragmentation related markers and test statistics. Patients showed trends for an overall increase in the amount of awakenings (unadjusted *p* = .053) and for a higher number of stage transitions (unadjusted *p* = .082) during nocturnal sleep when compared with controls. There was a nominally significant decrease in the overall sleep efficiency (unadjusted *p* = .043) in the patient group when compared with the controls. All of the abovementioned differences were no longer significant after correction for multiple comparisons. No other sleep architecture related parameters were significantly different across the groups or showed any interaction with age as a covariate (e.g., F_(1,79)_ = 0.26, *p* = 0.613 for the group × age interaction for SWS duration).

**Table 2.** Sleep architecture related markers in patients and controls. *P* values corresponding to one-sided *t*-tests and Mann–Whitney *U* tests are corrected for multiple comparisons (Benjamini–Hochberg correction). WASO = wake after sleep onset, SWS = slow-wave sleep; + significant before at *p* < 0.05, but not significant after FDR correction.

|  | Patient<br>(*N* = 47) |  | CTL<br>(*N* = 36) |  | Independent samples *t*-test /<br>Mann – Whitney *U* test |  |
| --- | --- | --- | --- | --- | --- | --- |
|  | Mean | SD | Mean | SD | *t*<sub>81</sub> or *U*<br>value | *p* value |
| Sleep latency (min) | 20.4 | 29.9 | 14.6 | 9.7 | 742.5 | .413 |
| Sleep efficiency (%) | 88.9 | 7.1 | 90.8 | 5.5 | 1033 | .318+ |
| Awakenings (#) | 22.8 | 8.3 | 20.1 | 6.6 | -1.636 | .318+ |
| WASO (min) | 29.3 | 28.1 | 24.1 | 19.3 | 738 | .413 |
| NREM stage 2 (min) | 190.4 | 45.9 | 185.4 | 44.1 | 776.5 | .526 |
| NREM stage 2 (%) | 47.2 | 7.7 | 47.2 | 7.8 | 877 | .748 |
| SWS (min) | 90.7 | 25.9 | 87.1 | 22.9 | -.671 | .748 |
| SWS (%) | 23.1 | 6.9 | 22.7 | 5.5 | -.331 | .748 |
| REM (min) | 113.9 | 31.4 | 112.5 | 36.4 | -.181 | .734 |
| REM (%) | 28.5 | 6.3 | 29.3 | 8.7 | 885.5 | .748 |
| Stage transitions (#) | 76.7 | 21.8 | 70.4 | 18.2 | -1.405 | .328 |
| SWS stage transitions<br>(#) | 7.4 | 2.8 | 7.8 | 2.7 | .663 | .748 |

### Overnight ascending gradient

Group differences in the overnight dynamicity of NREM sleep were investigated by an ANCOVA. Neither the group nor the effect of age were significant on any of our analyzed channels (Group: F7-O1 – F_1,74_ = .0136, *p* = .908; F8-O2 – F_1,68_ = 0.90, *p* = .346; Age: F7-O1 – F_1,74_ = 2.31, *p* = .133; F8-O2 – F_1,68_ = .03, *p* = .852). Nonetheless, there was a significant group × age interaction for both the F7-O1 (F_1,74_ = 6.71, *p* = .012) and F8-O2 (F_1,68_ = 4.13, *p* = .046) channels (see Figure 3), showing the expected flattening of overnight NREM slopes with age in the control group, and a fairly stable if not slightly increasing overnight ascending gradient with age in the patient group.

**Figure 3.**
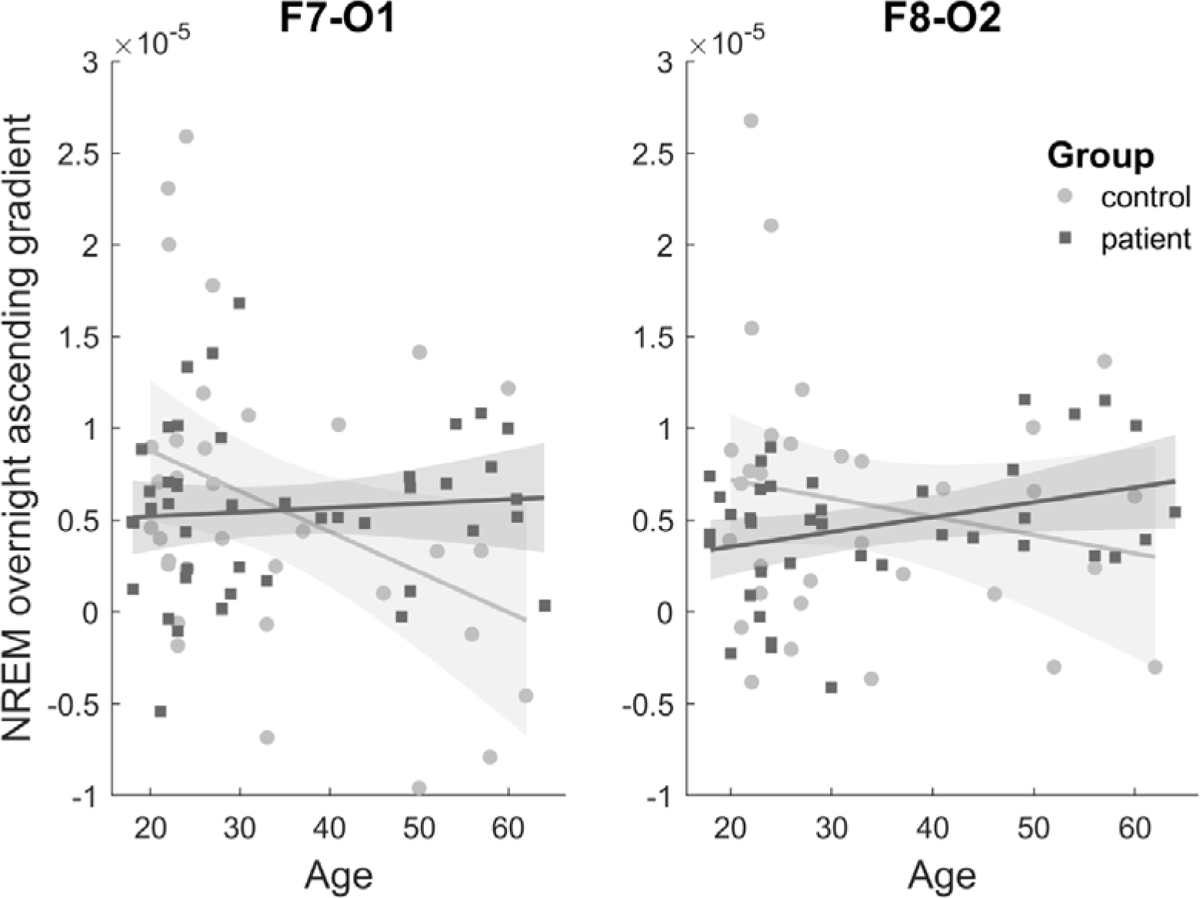
Age-related group differences in the NREM overnight ascending gradient on channels F7-O1 and F8-O2. Light gray circles represent the control group and dark gray squares the patient group. Lines indicate the fit of the generalized linear models with 95% confidence intervals.

### Heart rate and heart rate variability

First, a 2 × 3 rmANOVA was performed to examine differences in heart rate, where the group (control, patient) was the between-subject factor and the phase (pre-sleep wakefulness, sleep, post-sleep wakefulness) the within-subject factor. Here, we observed a significant effect of phase (F_2_ = 3.67, *p* = .03), no effect of group (F_1_ = 1.24, *p* = .27), and crucially, a significant group × phase interaction (F_1,2_ = 3.12, *p* = .05) that was driven by robust differences in mean heart rate in pre-sleep wakefulness (higher for patients) that became less pronounced in sleep and post-sleep wakefulness (see Figure 4a). This effect was not confounded by activity, because although activity also has a significant effect of phase (not of group) and group × phase interaction, this was in the other direction with robust differences in post-sleep wakefulness (see Supplementary Figure S4). Zooming in on the sleep stages and wake after sleep onset (WASO) with an explorative 2 × 4 rmANOVA did not reveal any significant main effects or interaction, with marginally increased heart rate values in patients throughout sleep stages and WASO (see Figure 4b). For RMSSD, neither the main effects (F_1,81_ < .09, *p* > .76, η*_p_*^2^ < .001) nor the interaction (F_1,81_ = .05, *p* = .82, η*_p_*^2^ < .001) was significant.

**Figure 4.**
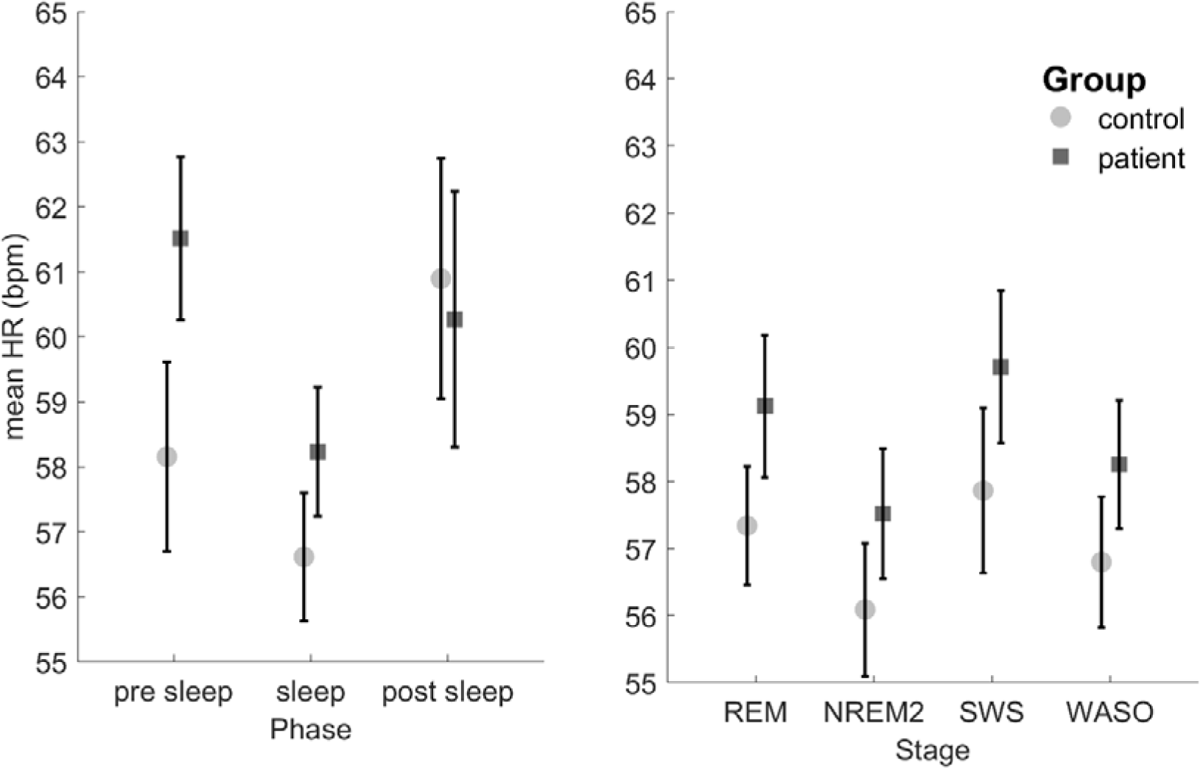
Mean heart rate (HR) differences between controls and patients. (a) Mean heart rate during pre-sleep wakefulness (up to 5 min), sleep, and post-sleep wakefulness (up to 5 min). (b) Mean heart rate during different sleep stages during the night. Light gray dots represent control participants, dark gray squares represent patients, and whiskers represent the standard error of the mean. Heart rates are in beats per minutes (bpm). REM = rapid-eye movement, NREM2 = non rapid-eye movement stage 2, SWS = slow-wave sleep, WASO = wake after sleep onset

## Discussion

We evaluated the utility of wearable headband EEG in a sample of healthy participants and unmedicated patients with stress-related mental disorders. To what extent could such headband (and wristwatch) wearables serve as measurement devices for sleep depth and fragmentation, sleep disorders, and transdiagnostic factors related to affective symptomatology such as cortical hyperarousal?

First, most of the participating patients seemed to manage well wearing the headband EEG for at least two nights, most for around two to nine nights. This might be because they had mild to moderate symptom intensities, and inpatients with more severe symptomatology may react differently. However, the headbands were better accepted than we anticipated at the start. Second, the data quality of the headband EEG surpassed our expectations by providing data of sufficient quality on a majority of nights in a majority of patients, not only for macrostructural analyses but also for more fine-grained spectral analyses of multiple epochs (robust to a rigorous artifact correction procedure). Third, our macrostructural results are largely in line with the scientific literature [26], that is, there were no large effects and only a few small to moderate nominally significant differences for macrostructural variables relating to sleep efficiency. This appears trivial, but suggests that sleep researchers have not missed any large effects in the last few decades due to potential context effects of the sleep [27,28] laboratory; instead, focusing on the second night as is typical in psychiatric sleep research seems to be a valid methodological solution. Severe first-night effects might still be relevant for more specific populations such as individuals with insomnia [27] and/or PTSD [29], who for instance report fewer nightmares in the sleep laboratory, potentially representing an expression of reduced sleep-related anxiety in the sleep laboratory.

Our spectral analyses revealed that the ratio of low versus high frequency power, characterized by the beta of the linear fit of the log-transformed 1/*f* power slope, did not differ between the groups. Instead, the age-related decline in this ratio [24] associated with reduced slow-wave power and sleep amount declined less in the patient sample. Patients had an apparent reduced ratio (less low-frequency, more high-frequency power) at a younger age than healthy participants and less of a decline with age. However, these observations are cross-sectional in nature and warrant replication in longitudinal data.

For our spectral analyses, we decided not to focus on analyzing the relevant frequency bands separately, because of false positive inflation and because the beta of the linear fit incorporated more relevant information than, for example, analyzing delta power alone. However, our results are in line with reported slow-wave power in the sleep of individuals with stress-related mental disorders, specifically with PTSD and depression [26]. We restricted our analyses to NREM sleep because REM sleep artifact detection requires more attention and clearer separation in the EEG traces of what is an eye movement and what is not. This calls for the development of more specific REM detection algorithms that are trained on frontal EEG electrodes with simultaneous electrooculography and can then be transferred to such data. For now, this restricts our spectral analyses to NREM, whereas REM sleep instability variables could be of particular relevance to this population [30].

This also raises the question to what extent our patients experienced insomnia, which is a common symptom of depression, with around 20% of people with depression having an Insomnia Severity Index score of 15 or above [31]. This is associated with increased depression severity, anxiety symptoms, and reduced activity levels [32]. However, we could not formally diagnose our participants with insomnia on the basis of an interview on their subjective reports. Naturally, we do have the actual data of sleep latency and WASO, the objective insomnia measures, and analyzed those (e.g., there were 10 participants with a sleep latency of 30 minutes or more in at least one night), but the literature suggests a great discrepancy between objective and subjective insomnia variables that prevents estimation of the prevalence of insomnia on the basis of objective sleep variables [33]. We believe, however, that sleep wearables such as headbands and wristwatches that combine actigraphy with heart rate will allow a more objective monitoring of sleep latency and WASO and have the potential to bring insomnia diagnostics back to sleep medicine. Moreover, we observed that three healthy controls and five patients had a median of six or more SWS-to-wake transitions per night, which could be a sign of more organic sleep problems relating to sleep apnea or periodic limb movements. This suggests that in a psychiatric patient sample, with depressed inpatients having surprisingly high prevalence of sleep apnea [34], headbands in combination with breathing-related wearables [35] might be used as an initial screening device for optimized decisions on who is referred to full polysomnography in the clinical sleep laboratory.

This then brings us to our heart rate and activity analyses, which revealed a group × time interaction for mean heart rate. One explanation would be an upregulation of sympathetic compared with parasympathetic activity before falling asleep, perhaps in apprehension of the upcoming night (related to a fear of sleep [36] in PTSD, but a milder variant in stressed participants), which reduces during but remains present throughout sleep. An alternative explanation is that because there appeared marginally increased differences throughout the sleep stages, this might reflect a more general reduced physical condition in the patients, which would be in line with the literature on reduced activity in people with stress-related mental disorders, something that can be reliably detected with actigraphy [37]. However, this explanation cannot account for a decline in the differences at post-sleep, which seems to point more in the direction of a homeostatic process. The heart rate variability measures did not reveal any effects of interest, but our metric, the most commonly used – the RMSSD – is highly susceptible to noise in the data. Peaks in the PPG have a lower kurtosis than peaks in the electrocardiograph (ECG), easily adding some additional milliseconds of variation to the peak-to-peak differences, and flattened peaks also occur and could lead to a missing peak of additional variation in the peak timing. This is also why pulse rate variability from PPG is a different metric than heart rate variability from the ECG [38], and there is still a debate about what is the most artifact-resistant way to analyze heart rate variability in sleep. RMSSD is probably not going to win it.

Further limitations include the restriction of spectral analyses to NREM sleep until REM algorithms have been properly trained and openly shared, the cross-sectional nature of the data in the light of the group × age interactions, and the lack of formal insomnia diagnoses based on subjective data. We could confirm the mental disorders of patients, and the absence of disorders in healthy participants in our sample, but the sample itself was too small, and the overlap among mental disorders too high, to analyze the data for subgroups (e.g., major depressive disorder (MDD) without an anxiety disorder, MDD with an anxiety disorder, anxiety disorder without MDD). Finally, we do not know the ground truth of true sleep in our sample, as we do not have 1–2 nights with polysomnography in the sleep laboratory. Although the accuracy of the headband EEG compared with our raters was high, a recent study reported that the Dreem headband appears to underestimate wakefulness and overestimate NREM 1/NREM 2 [21]. Given that sleep continuity related variables showed nominally significant group differences in our work, such a systematic bias in the detection of wakefulness versus light sleep could be a confound. However, we do not claim any epidemiological ground truth in the amount of hours slept or WASO minutes, but instead focused on group differences that should have the same systematic bias given the similarity in sleep patterns. However, whether the systematic bias is actually the same for both groups is still an assumption, and further research should address the question of to what extent differences in activity might affect detection of wakefulness in both headband-based and wrist-based wearables.

In conclusion, the headbands were accepted well by people with stress-related mental disorders, and the data quality was sufficient for macro- and microstructural analyses of a majority of nights in a majority of participants. As in previous research, we did not observe large macrostructural between-group effects except for those related to fragmented sleep. More temporally fine-grained analyses revealed differences in age-related changes in the low versus high frequency power ratio and in nightly activity and mean heart rate. Our results imply that wearable devices that record EEG and/or PPG can be used over multiple nights to assess sleep depth and fragmentation, cortical hyperarousal, and sympathetic drive throughout the sleep–wake cycle in a sample of people with stress-related mental disorders.

## Data Availability

The data underlying this article will be shared on reasonable request to the corresponding author.

## Acknowledgments

BeCOME working group includes Elisabeth B. Binder, Angelika Erhardt, Susanne Lucae, Philipp G. Saemann, Norma C. Grandi, Tamara Namendorf, Michael Czisch, Immanuel Elbau, Laura Leuchs, Anna Katharine Brem, Leonhard Schilbach, Julia Fietz, Sanja Ilić-Ćoćić, Julius Ziebula, Iven-Alex von Mücke-Heim, Yeho Kim and Julius Pape.

We would like to thank Stephanie Alam, Julia-Carolin Albrecht, Anastasia Bauer, Anja Betz, Miriam El-Mahdi, Gertrud Ernst-Jansen, Carolin Haas, Karin Hofer, Lisa Kammholz, Elisabeth Kappelmann, Sophia Koch, Alexandra Kocsis, Anna Lorenz, Rebecca Meissner, Jessie Osterhaus, Lisbeth Pirn and Linda Schuster for their help with data collection, study management, the recruitment, and screening of BeCOME participants. The authors’ special thanks go to all study participants for their participation in the BeCOME study.

## Data Availability Statement

The data underlying this article will be shared on reasonable request to the corresponding author.

