## Supplementary Material for "The utility of wearable headband electroencephalography and pulse photoplethysmography to assess cortical hyperarousal in individuals with stress-related mental disorders"

**Supplementary Materials**

**Supplementary Table S1**

| **Metric / Stage** | **SWS** | **NREM2** | **NREM1** | **REM** | **Wake** |
| --- | --- | --- | --- | --- | --- |
| **Sensitivity** | .98 | .94 | .65 | .93 | .77 |
| **Specificity** | .99 | .95 | .99 | .96 | .99 |
| **Precision** | .96 | .93 | .59 | .89 | .92 |
| **Overall Accuracy** | .92 | | | | |

*Evaluation of the automated Dreem scoring algorithm compared to manually re-scored epochs.*

**Supplementary Figure S1**

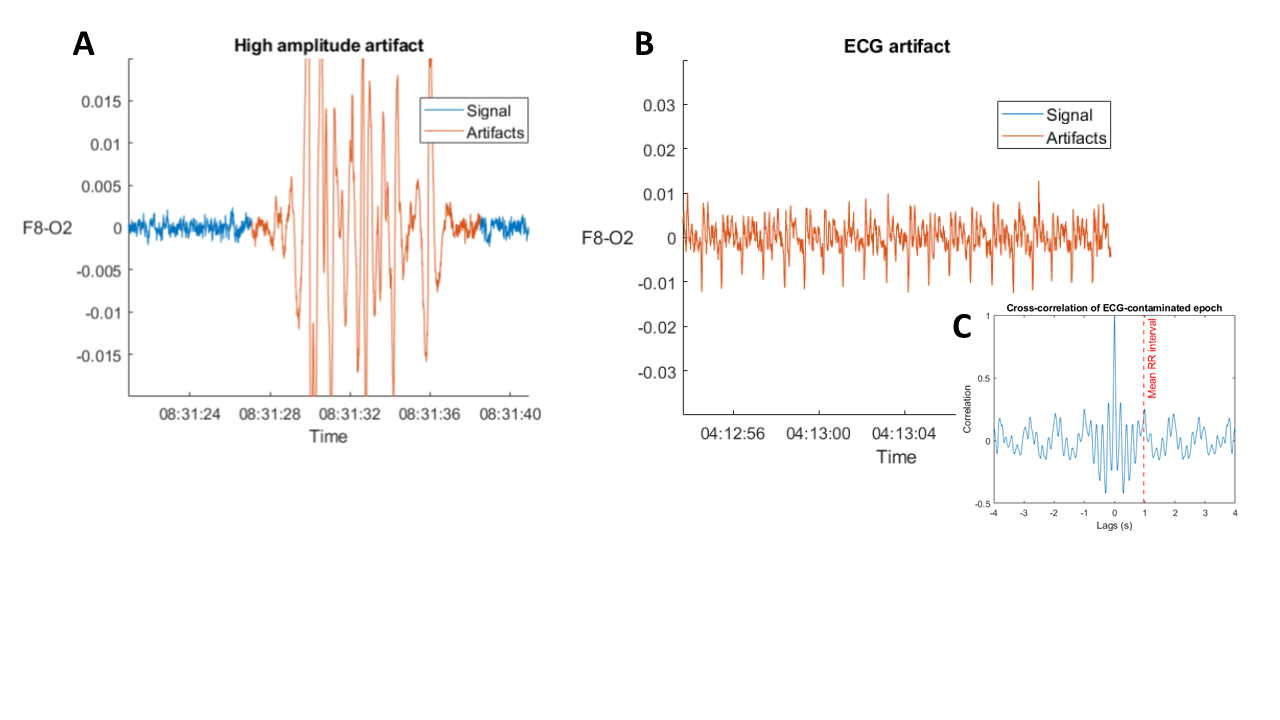

**Figure S1***. Examples of detected artifacts from each of the filters. (A) Detection made by the above-threshold filter. (B) Detection made by the cardiac artifacts filter. Notice the periodicity of the signal and the equally spaced peaks. (C) Cross-correlation from the signal represented in B. In red dashed line: mean RR interval for the epoch, computed from the PPG signal. The cross-correlation shows peaks at lags which are multiples of the mean RR interval.*

**Supplementary Figure S2**

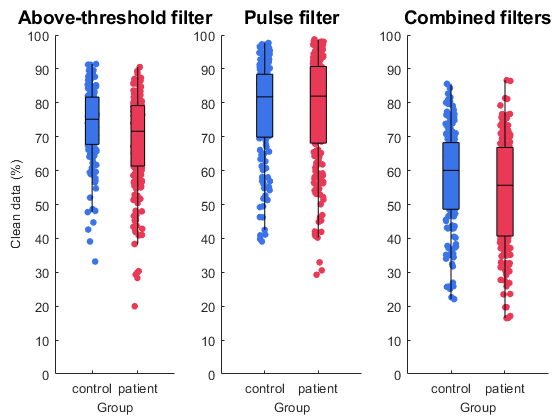

**Figure S2**. *Percentage of signal classified as clean by each filter and the combined use of them.*

**Supplementary Figure S3**

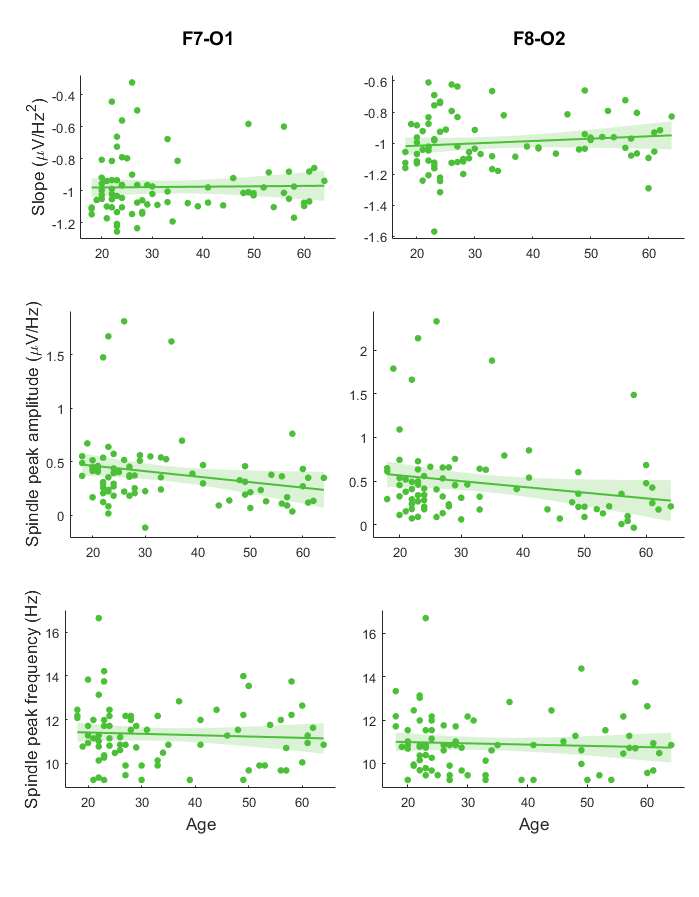
**Figure S3***. Age-related changes in NREM overall slope, spectral peak amplitude and spectral peak frequency in the sleep spindle frequency range for the two long-range channels. Age related changes in NREM spectral slope, spindle amplitude and frequency were calculated with Pearson’s correlation coefficient.*

**Supplementary Figure S4**

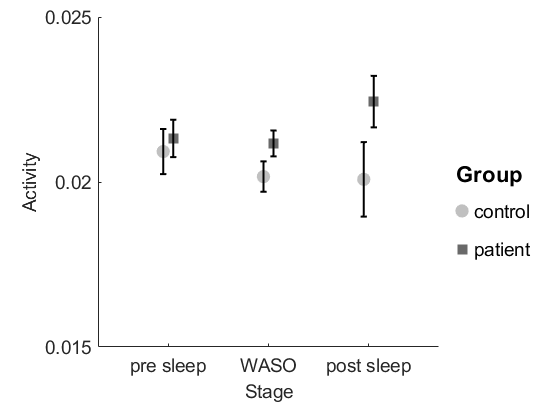

**Figure S4.** *Activity measurements based on the built-in EEG-headband accelerometer values represented for the two groups separately during pre-sleep, during sleep and after sleep. Pre-sleep phases were max. 5 min awake period right before stable NREM2 sleep sets in and post-sleep phases were defined as max. 5 minutes right after the last awakening. Activity values were only taken from periods when the participants were lying in bed.*
